# Causal Relationship Between Gut Microbiota and Androgenic Alopecia: A Mendelian Randomization Analysis

**DOI:** 10.1101/2025.04.10.25325503

**Authors:** Shuning Liu, Debin Xu

**Affiliations:** School of Marxism, Changchun University of Chinese Medicine, Changchun 130117, China

**Keywords:** Gut Microbiota, Androgenetic Alopecia, Causal Relationship, Mendelian Randomization

## Abstract

**Background:** Androgenic alopecia (AGA) is a common condition influenced by genetic and hormonal factors, with emerging evidence linking it to gut microbiota. Dysbiosis may affect AGA through immune regulation, androgen metabolism, and systemic pathways. Using Mendelian randomization (MR), this study investigates the causal relationship between gut microbiota and AGA, aiming to uncover potential mechanisms and therapeutic targets.

**Methods:** The study selected six types of gut microbiota-related genetic loci associated with GM from summary data of genome-wide association studies (GWAS), which served as instrumental variables. MR analysis was conducted utilizing inverse variance weighting (IVW), MR-Egger regression, and weighted median approaches. Sensitivity analyses were performed through heterogeneity tests, pleiotropy tests, and leave-one-out analyses, while odds ratios and 95% confidence intervals were used to assess the potential causal relationship between GM and AGA.

**Results:** The IVW analysis revealed that several taxa may serve as protective factors against AGA, including the Oxalobacteraceae (OR=0.957, 95% CI 0.919-0.996, P=0.033), Paraprevotella (OR=0.953, 95% CI 0.909-0.999, P=0.047), Eubacteriumventriosumgroup (OR=0.931, 95%CI 0.869-0.997, P=0.043), LachnospiraceaeUCG008 (OR=0.937, 95%CI 0.892-0.985, P=0.011), and Clostridiales (OR=0.917, 95%CI 0.853-0.986, P=0.019). Conversely, the Coriobacteriaceae (OR=1.103, 95%CI 1.003-1.213, P=0.041) may be a potential risk factor for AGA. Heterogeneity testing indicated no significant heterogeneity in results (P>0.05), and the intercept term from MR-Egger regression ruled out the presence of horizontal pleiotropy. The leave-one-out test results consistently fell on one side of the null line, indicating good robustness in MR analysis. Funnel plot analysis demonstrated that the distribution of all SNPs was approximately symmetrical, elucidating that potential confounding factors have relatively small impact on causality.

**Conclusion:** This study suggests a potential causal relationship between gut microbiota and AGA, highlighting potential pathways and opening opportunities for microbiota-based therapeutic strategies.

## 1 Introduction

Androgenetic alopecia (AGA) is one of the most prevalent types of hair loss in clinical practice, characterized by a significant genetic predisposition and a close correlation with androgen levels in the human body [1]. In China, over 250 million individuals are facing issues related to hair loss, with the onset age of this condition showing a declining trend, which has had a substantial impact on personal image [2]. AGA primarily results from excessive secretion of androgens in the scalp or heightened sensitivity to these hormones. The hallmark features of this condition include miniaturization of hair follicle cells in the frontal and crown areas as well as a reduction in the growth phase duration of dermal cells. This leads to an increasing density of damaged hair follicles and subsequently causes gradual hair thinning [3, 4]. The gut microbiota (GM) refers to the vast collection of microorganisms residing within the gastrointestinal tract. These microbes play crucial roles by absorbing nutrients that stimulate intestinal cells and participating in immune responses, thereby influencing the host’s immune system. With ongoing research into the Gut-Skin Axis, it has become evident that GM plays an important role in maintaining systemic immune homeostasis [5]. Due to bidirectional communication between GM and skin health, various pathways such as immune modulation and transfer of strains or metabolic products can affect skin health [6-8]. MR is a research methodology that utilizes randomly assigned genetic variations within populations as instrumental variables. By establishing associations between these instrumental variables, exposure factors, and outcomes, MR aims to reveal potential causal relationships between exposure factors and outcomes [9]. Given that genetic variations are randomly allocated during meiosis—once alleles are formed they remain unaffected by subsequent confounding factors—MR can simulate effects akin to those observed in randomized controlled tri-

## 2 Materials and Methods

### 2.1 Data Sources

The GWAS data was obtained from the MiBioGen consortium, which encompasses a total of 18,340 individuals across 24 distinct cohorts. For genetic data related to AGA, we utilized the GWAS dataset published by UK Biobank in 2021, comprising 66,172 cases and 140,864 control individuals, amounting to a total of 207,036 subjects. Both datasets are derived from populations of European descent and do not differentiate by sex. All data are from publicly available datasets, so no additional ethical approval is required.

### 2.2 Instrumental Variable

The instrumental variable in MR studies must satisfy three assumptions: (1) Relevance assumption: There is a robust strong correlation between the instrumental variable and the exposure factor; (2) Independence Assumption: The instrumental variables remain independent of confounding factors affecting the “exposure-outcome” relationship; (3) Exclusivity Assumption: Genetic variations influence outcomes solely through exposure factors without any alternative pathways impacting the outcome [12]. SNPs significantly associated with GM (P<1×10^−5^) were screened while setting (r^2^<0.001 and kb=10,000) to exclude linkage disequilibrium (LD). Using the PhenoScanner database (P<5×10^−8^; r^2^>0.8), SNPs directly correlated with confounding factors were excluded based on F-statistic values to remove weak instrumental variables als [10, 11]. This study employs MR methods to analyze causal links between GM and AGA with an aim to provide reference for future research (Figure 1). (F<10) [13].

**Figure 1.**
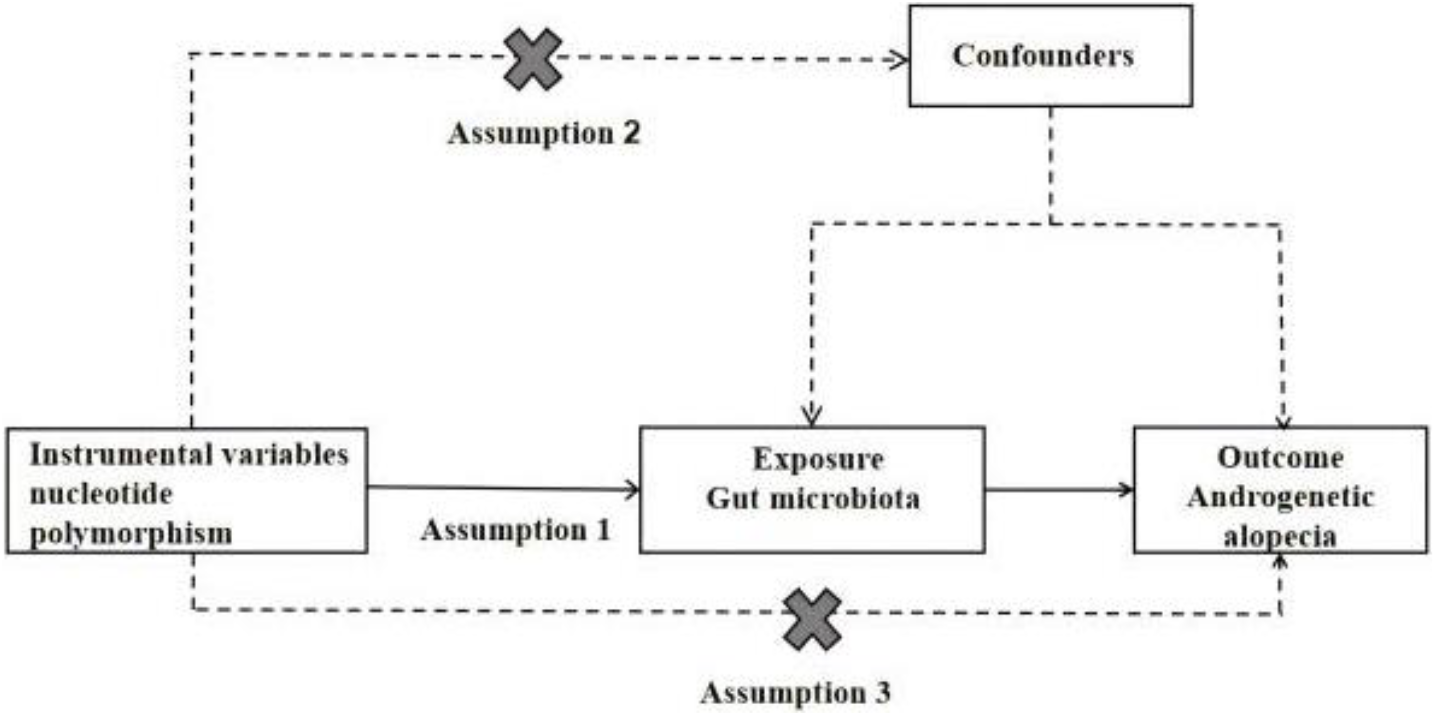
Schematic diagram of MR analysis

### 2.3 MR Analysis

MR analysis primarily employed Inverse Variance Weighting (IVW), Weighted Median, and MR-Egger regression methods [14]. When selected SNPs serve as effective instruments for IV analysis, IVW yields the most precise causal effect estimates [15]. The advantage of Weighted Median lies in its ability to provide consistent causal effect estimates even when some invalid instrumental variables are present [16]. The MR-Egger regression method accounts for potential heterogeneity among instrumental variables and provides adjusted causal effect estimates capable of detecting and correcting pleiotropic bias among instruments [17]. Ultimately, results from IVW will be fitted alongside those from Weighted Median and MR-Egger regression methods [18], with outcomes assessed using Odds Ratios (OR) and corresponding 95% Confidence Intervals (CI) to evaluate potential causal associations.

### 2.4 MR Sensitivity Analysis

Cochran’s Q test was employed to assess whether there exists significant variability between two sets of results; a P-value less than 0.05 indicates notable differences [19]. To detect horizontal pleiotropy via MR-Egger intercept method: if there is a significant difference between the intercept term and zero along with a P-value for the regression intercept below 0.05, it suggests evidence of horizontal pleiotropy [20]. Additionally, Leave-one-out testing was conducted to ascertain whether individual SNPs exert an influence on MR results.

## 3 Results

### 3.1 Instrumental Variables

After excluding confounding factors and palindromic SNPs, a total of 69 SNPs were included in the final analysis. The F-statistics for all selected SNPs in this study exceeded 10, indicating that the results are reliable and there is no concern regarding weak instrument bias. Following the exclusion of potential confounders and palindromic SNPs, we confirmed the inclusion of 69 SNPs for further analysis. The F-statistics associated with these selected SNPs consistently surpassed 10, reinforcing the reliability of our findings while eliminating any possibility of weak instrument bias.

### 3.2 MR Analyses

According to the IVW results, increased abundance of certain bacterial taxa was found to be associated with a reduced risk of AGA: the Oxalobacteraceae (OR=0.957, 95% CI 0.919-0.996, P=0.033), Paraprevotella (OR=0.953, 95% CI 0.909-0.999, P=0.047), Eubacteriumventriosum group (OR=0.931, 95%CI 0.869-0.997, P=0.043), LachnospiraceaeUCG008 (OR=0.937, 95%CI 0.892-0.985, P=0.011), and Clostridiales (OR=0.917, 95%CI 0.853-0.986, P=0.019) may serve as protective factors against AGA risk. Conversely, an increase in abundance of the Coriobacteriaceae was linked to an elevated risk for AGA (OR=1.103, 95%CI 1.003-1.213, P=0.041), suggesting its potential role as a risk factor for this condition (Table 1). The associations between GM and AGA were revealed through IVW analysis along with MR-Egger regression and weighted median approaches, where colored lines represent fitting results from these three methodologies (Figure 2).

**Table 1.** MR Analysis Results.

| Conclusion | Exposure | Method | OR (CI95%) | $\beta$ | SE | P |
| --- | --- | --- | --- | --- | --- | --- |
| Androgenetic alopecia | Oxalobacteraceae | IVW | 0.957 (0.919- 0.996) | -0.043 | 0.02 | 0.033 |
|  |  | Weighted Median | 0.955 (0.906- 1.006) | -0.045 | 0.026 | 0.045 |
|  |  | MR-Egger | 0.999 (0.848- 1.177) | -0.001 | 0.083 | 0.099 |
|  | Paraprevotella | IVW | 0.953 (0.909- 0.999) | -0.047 | 0.024 | 0.047 |
|  |  | Weighted Median | 0.976 (0.915- 1.041) | -0.023 | 0.032 | 0.047 |
|  |  | MR-Egger | 0.888 (0.758- 1.042) | -0.117 | 0.081 | 0.174 |
|  | Eubacterium ventriosum group | IVW | 0.931 (0.869- 0.997) | -0.070 | 0.035 | 0.043 |
|  |  | Weighted Median | 0.924 (0.84- 1.017) | -0.078 | 0.048 | 0.011 |
|  |  | MR-Egger | 1.136 (0.845- 1.527) | 0.128 | 0.150 | 0.413 |
|  | LachnospiraceaeUCG008 | IVW | 0.937 (0.892- 0.985) | -0.064 | 0.025 | 0.011 |
|  |  | Weighted Median | 0.949 (0.887- 1.015) | -0.051 | 0.034 | 0.013 |
|  |  | MR-Egger | 0.977 (0.759- 1.259) | -0.022 | 0.128 | 0.866 |
|  | Clostridiales | IVW | 0.917 (0.853-0.986) | -0.085 | 0.036 | 0.019 |
|  |  | Weighted Median | 0.909 (0.826- 1.001) | -0.094 | 0.048 | 0.032 |
|  |  | MR-Egger | 1.079 (0.768- 1.516) | 0.076 | 0.173 | 0.665 |
|  | Coriobacteriaceae | IVW | 1.103 (1.003- 1.213) | 0.098 | 0.048 | 0.041 |
|  |  | Weighted Median | 1.082 (0.975- 1.201) | 0.079 | 0.052 | 0.042 |
|  |  | MR-Egger | 1.135 (0.769- 1.675) | 0.127 | 0.198 | 0.533 |

**Figure 2.**
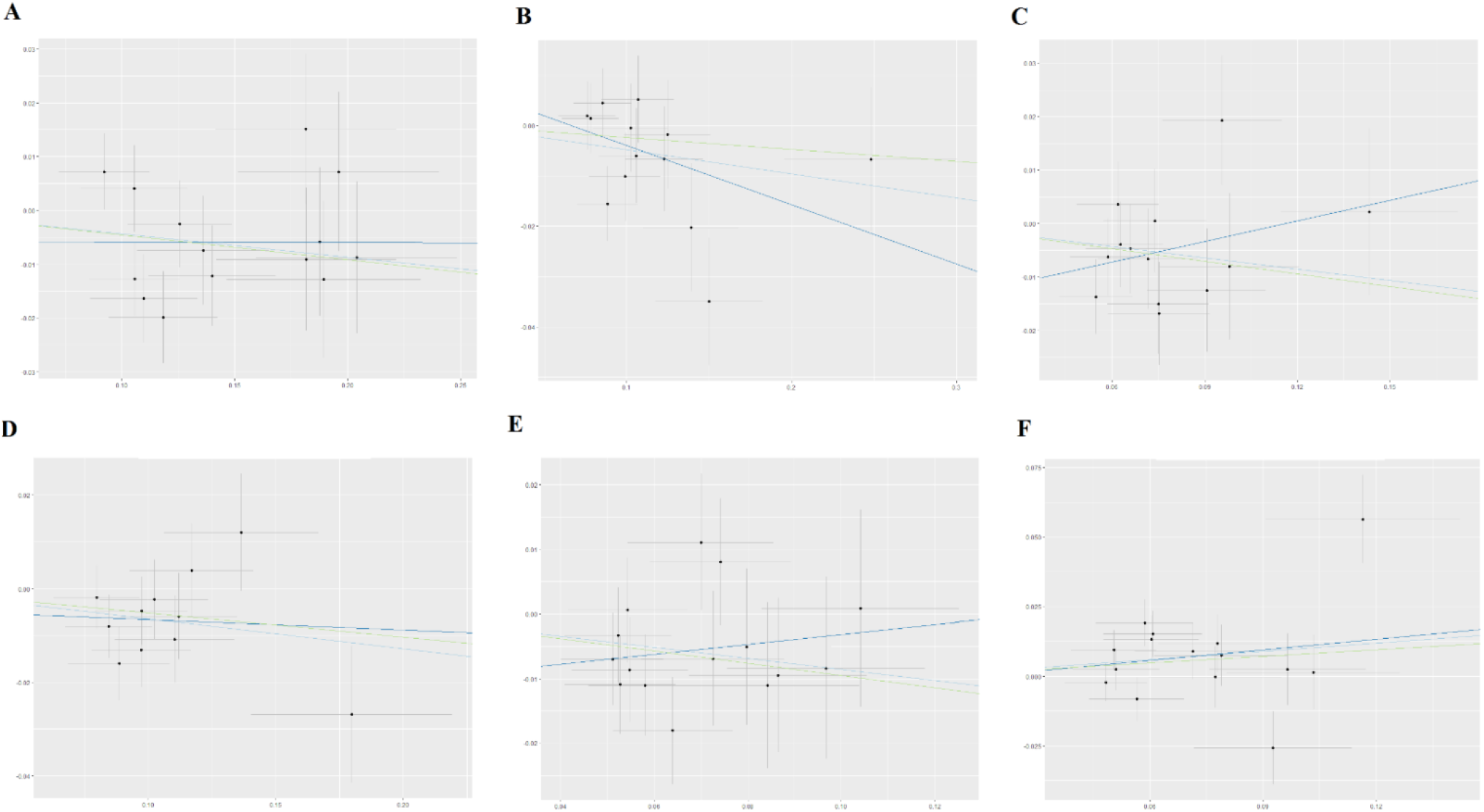
Scatter Plot of MR Results (A. Oxalobacteraceae; B. Paraprevotella; C. Eubacteriumventriosum group; D. LachnospiraceaeUCG008; E. Clostridiales; F. Coriobacteriaceae)

### 3.3 MR Sensitivity Analyses

Cochran’s Q test assessed heterogeneity within data results from both IVW and MR-Egger regression analyses; no evidence of heterogeneity was detected (P> 0.05). To evaluate horizontal pleiotropy, we examined whether there was deviation between the intercept term obtained from MR-Egger regression and zero; our findings indicated absence thereof (P > 0.05) (Table 2). Funnel plot analysis demonstrated that all SNP distributions were approximately symmetrical, confirming robustness in causal associations. Leave-one-out tests evaluated each individual SNP’s impact on overall causal relationships; upon removing one SNP at a time, remaining ones consistently fell on one side relative to null effect line without identifying any single influential outlier which suggests that our MR analytical outcomes are not driven by any particular SNP but rather exhibit substantial robustness across analyses performed. By assessing distances between intercept terms derived from MR-Egger regressions compared against zero points enables us ascertain presence or absence concerning horizontal pleiotropy; analyses yielded conclusive evidence negating such effects (P>0.05) (Table 2). Furthermore, funnel plots illustrated near-symmetrical distribution among all considered variants affirming stability inherent within observed causal links (Figure 3). Lastly leave-one-out assessments gauged influence exerted by every variant upon collective causation revealing consistent positioning beyond threshold delineating non-effectiveness post removal thus underscoring resilience exhibited throughout conducted investigations thereby validating integrity surrounding resultant conclusions drawn herein (Figure 4).

**Table 2.** Results of Sensitivity Analysis.

| Microbiota | Cochran's Q test |  | MR-Egger regression |  |
| --- | --- | --- | --- | --- |
|  | Q | P | Egger-intercept | P |
| Oxalobacteraceae | 14.545 | 0.336 | -0.005 | 0.605 |
| Paraprevotella | 13.387 | 0.341 | 0.007 | 0.385 |
| Eubacteriumventriosum group | 11.473 | 0.488 | -0.014 | 0.202 |
| LachnospiraceaeUCG008 | 8.417 | 0.588 | -0.004 | 0.748 |
| Clostridiales | 9.738 | 0.781 | -0.011 | 0.354 |
| Coriobacteriaceae | 25.522 | 0.029 | -0.001 | 0.885 |

**Figure 3.**
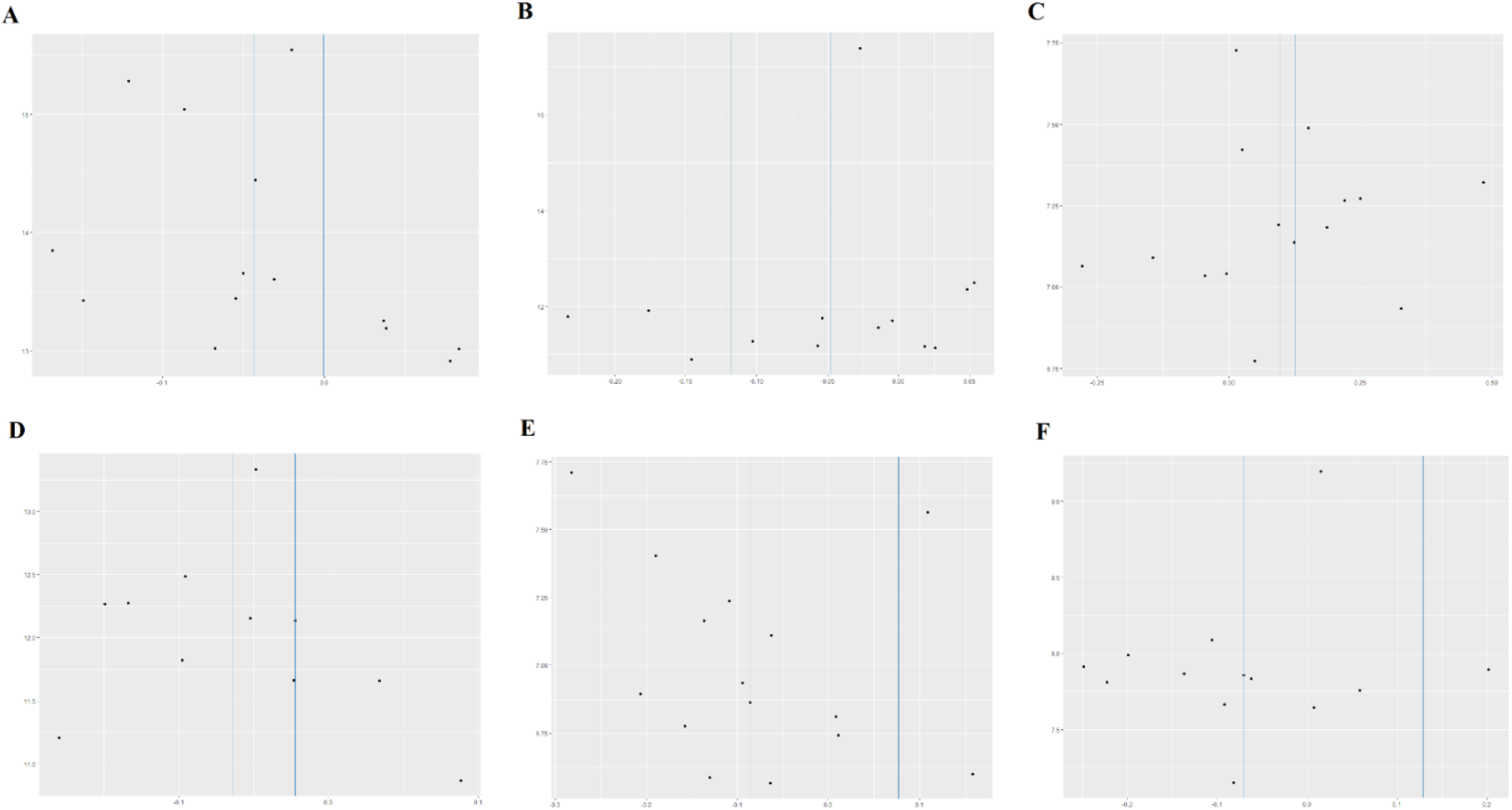
Heterogeneity Funnel Plot (A. Oxalobacteraceae; B. Paraprevotella; C. Eubacteriumventriosum group; D. LachnospiraceaeUCG008.; E. Clostridiales.; F. Coriobacteriaceae)

**Figure 4.**
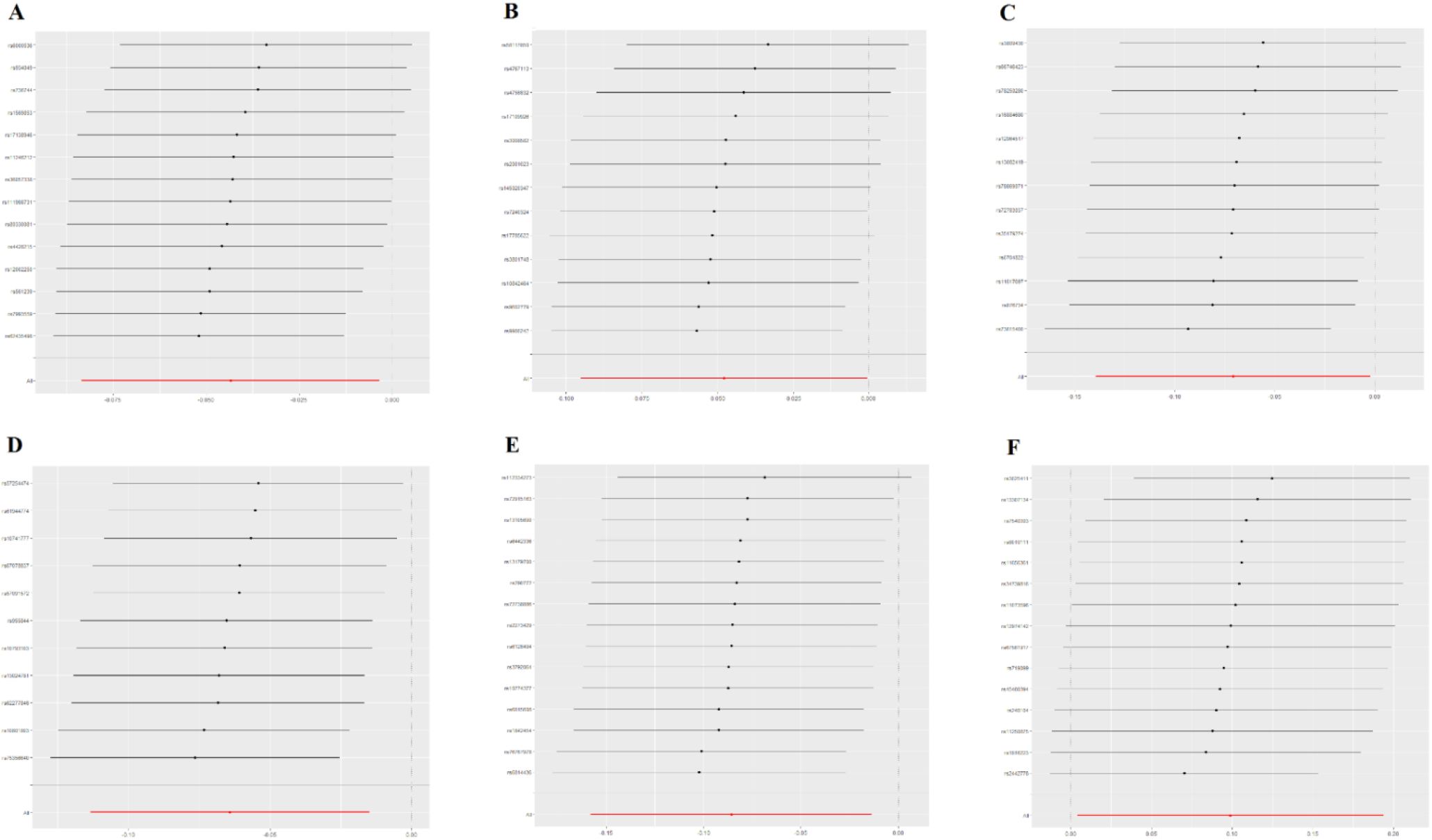
Leave-one-out Analysis Results (A. Oxalobacteraceae; B. Paraprevotella; C. Eubacteriumventriosum group; D. LachnospiraceaeUCG008; E. Clostridiales; F. Coriobacteriaceae)

## 4 Discussion

This study analyzes the relationship between GM and AGA using MR methods. The findings indicate that five specific GM groups may serve as protective factors against AGA: Oxalobacteraceae (OR=0.957, 95% CI 0.919-0.996, P=0.033), Paraprevotella (OR=0.953, 95% CI 0.909-0.999, P=0.047), Eubacteriumventriosumgroup (OR=0.931, 95%CI 0.869-0.997, P=0.043), LachnospiraceaeUCG008 (OR=0.937, 95%CI 0.892-0.985, P=0.011), and Clostridiales (OR=0.917, 95%CI 0.853-0.986, P=0.019). Conversely, the Coriobacteriaceae (OR=1.103, 95%CI 1.003-1.213, P=0.041) may be a potential risk factor for AGA. The research suggests a bidirectional association between GM and AGA; specifically, levels of the enzyme 5α-reductase in hair follicles from balding areas are significantly higher than those in non-balding regions [21]. This enzyme facilitates the conversion of testosterone to dihydrotestosterone within tissues—leading to follicular miniaturization and subsequent abnormal hair growth patterns over time which can result in gradual follicle atrophy until complete loss occurs [22]. The gastrointestinal tract is one of the largest epithelial tissues in the human body with a surface area ranging from approximately 250 to 400 square meters housing over 500 species of bacteria. The predominant bacterial phyla include Bacteroidetes and Firmicutes while Proteobacteria Actinobacteria and Fusobacteriaphyla are commonly found in these areas [23]. Rebello D et al. conducted an experiment wherein they introduced Clostridium difficile into patients’ intestines via fecal microbiota transplantation techniques, resulting in observable hair regrowth on various parts including scalp, facial, and armregions [24]. Additionally, Alkhalifah A et al. analyzed stool samples from patients with alopecia areata constructing predictive models for disease status. They discovered that models based on Subdoligranulum and Butyricicoccus could accurately predict patient conditions [25]. Moreover, the alteration of GM can interact with immune systems through harmful metabolites increasing pro-inflammatory cells, cytokines, and metabolites circulating into distal organs such as brain, lungs, and skin leading to inflammation states within specific extraintestinal organs [26]. The onset of skin diseases not only triggers changes within cutaneous microbiomes but also induces corresponding modifications among intestinal flora, suggesting possible bidirectional signaling pathways between skin and gut [27]. This pathway plays a crucial role in alleviating inflammation across extraintestinal organs while improving symptom associated with disease [28]. Liu C et al. identified alterations in GM structure among patients suffering from melasma as well as the biological effects offirmicutes with in microbial communities potentially influencing β-glucuronidase production alongside androgen synthesis or metabolism thereby contributing to melasma development [29]. In summary, these studies reveal a potential association between Clostridium, Methanobrevibacter, Rhodococcus, Oxalobacter, and Faecalibacterium with neuro inflammation and immune damage in AGA.

The study reveals associations between lifelong carriers of alleles related to AGA in the general population, distinguishing it from traditional randomized controlled trials. This allows for a comprehensive assessment of the role of GM in the prevention or treatment of AGA [30]. MR analysis mitigates interference from confounding factors and bidirectional causal relationships in association analyses while considering the long latency period of diseases and the impact of individuals’ lifetime exposure to risk factors, significantly reducing inherent biases present in observational studies. Causal effects were evaluated using inverse variance weighting, weighted median methods, and MR-Egger regression analysis, with sensitivity analyses further confirming the stability and reliability of results. The data for this MR study was sourced from populations of European descent, utilizing recent GWAS findings on both GM and AGA, which provided larger sample sizes and higher statistical power. However, this research has certain limitations. Firstly, GWAS data lacks detailed statistics regarding participant sex ratios, environmental factors, racial differences, and disease duration; thus complicating comparisons of causal effects across different subgroups. The presence of multiple etiologies and subtypes may also affect the precision of research outcomes. Secondly, this study exclusively included individuals from European ancestry without analyzing other ethnic groups; although this helps reduce population stratification bias, its applicability to other racial groups may be limited [31]. While this study confirms that GM has a protective effect against AGA, it does not explore potential mediating factors influencing these results. Interventions solely targeting GM may not provide optimal solutions for mitigating or treating AGA risk due to numerous other non-genetic factors, such as unemployment status, psychosocial influences, poverty levels, educational opportunities and access to healthcare services, which were not incorporated into this research nor obtainable through GWAS methodologies.

## 5 Conclusion

This study provides evidence for a potential causal relationship between gut microbiota composition and AGA. Dysbiosis may influence AGA through immune modulation, androgen metabolism, and systemic pathways. These findings highlight the importance of gut health in AGA pathogenesis and open new avenues for therapeutic intervention. Future research should explore the underlying mechanisms and evaluate the efficacy of microbiota-targeted therapies in preventing or treating AGA, offering a novel perspective on managing this widespread condition.

## Data Availability

All data produced in the present work are contained in the manuscript.

## Acknowledgments

The authors thank all participants and investigators who provided the GWAS data.

## Author Contributions

Shuning Liu: Investigation, Visualization, Software, Conceptualization, Formal analysis, Methodology, Project administration, Data curation, Resources, Supervision, Validation, Writing – original draft.

Debin Xu: Project administration, Supervision, Writing – review & editing.

Institutional Review Board Statement: All data are publicly available GWAS datasets, therefore no additional ethical approval was required.

## Conflicts of Interest

The authors declare no conflicts of interest.

## Ethics Declaration

All data produced in the present work are contained in the manuscript.

MiBioGen: https://mibiogen.gcc.rug.nl

UK Biobanks: https://www.ukbiobank.ac.uk:

https://www.ebi.ac.uk/gwas/studies/GCST90043616

## Notes

### Competing Interest Statement

The authors have declared no competing interest.

### Funding Statement

This study did not receive any funding.

### Author Declarations

The study used (or will use) ONLY openly available human data that were originally located at: MiBioGen: https://mibiogen.gcc.rug.nl UK Biobanks: https://www.ukbiobank.ac.uk: GWAS dataset: https://www.ebi.ac.uk/gwas/studies/GCST90043616

